# Keeping doors open: A cross-sectional survey of family physician practice patterns during COVID-19, needs, and intentions

**DOI:** 10.1101/2021.12.20.21267918

**Authors:** Tara Kiran, Rick Wang, Curtis Handford, Nadine Laraya, Azza Eissa, Pauline Pariser, Rebecca Brown, Cheryl Pedersen

**Affiliations:** Department of Family and Community Medicine, Temerty Faculty of Medicine, University of Toronto, Toronto, Ontario; MAP Centre for Urban Health Solutions, Li Ka Shing Knowledge Institute, St. Michael’s Hospital, Toronto, Canada; Department of Family and Community Medicine, St. Michael’s Hospital, Toronto, Ontario; Institute of Health Policy, Management and Evaluation, University of Toronto, Toronto, Ontario

**Author notes:** **Corresponding author:** Tara Kiran, MD, MSc, MAP Centre for Urban Health Solutions, St. Michael’s Hospital, Unity Health Toronto, 30 Bond St., Toronto, ON Canada M5B 1W8. **Author contributions** TK conceived of the study and designed it together with members of the study working group and Survey Research Unit (SRU). The SRU collected the data in collaboration with regional primary care collaborators. RW, CP, and the SRU team conducted the analysis. All authors helped interpret the data. TK drafted the manuscript and all authors critically reviewed it. All authors read and approved the final manuscript and agree to be accountable for all aspects of the work.

## Abstract

**Objective:** To determine the extent to which family physicians closed their doors altogether or for in-person visits during the pandemic, their future practice intentions, and related factors.

**Methods:** Between March and June 2021, we conducted a cross-sectional survey using email, fax, and phone of 1,186 family doctors practicing comprehensive family medicine in Toronto, Ontario. We asked about practice patterns in January 2021, use of virtual care, and practice intentions.

**Results:** Of the 1,016 (86%) that responded to the survey, 99.7% (1001/1004) indicated their practice was open in January 2021 with 94.8% (928/979) seeing patients in-person and 30.8% (264/856) providing in-person care to patients reporting COVID-19 symptoms. Respondents estimated spending 58.2% of clinical care time on phone visits and an additional 5.8% on video and 7.5% on email. 17.2% (77/447) were planning to close their current practice in the next five years. There was a higher proportion of physicians who worked alone in a clinic among those who did not see patients in-person (27.6% no vs 12.4% yes, p<0.05), did not see symptomatic patients (15.6% no vs 6.5 % yes, p<0.001), and those who planned to close their practice in the next 5 years (28.9% yes vs 13.9% no, p<0.01).

**Interpretation:** The vast majority of family physicians in Toronto were open to in-person care in January 2021 but almost one-fifth are considering closing their practice in the next five years. Policy-makers need to prepare for a growing family physician shortage and better understand factors that support recruitment and retention.

## Introduction

The COVID-19 pandemic has placed inordinate stress on primary care, the front door of our healthcare system. Most family physicians in Canada and the US are self-employed individuals running independent practices and were suddenly responsible for enacting numerous changes to keep themselves, their patients, and staff safe. To safely see patients in-person, family physicians needed to adopt a range of measures including personal protective equipment, improved ventilation, enhanced cleaning, passive and active symptom screening, physical distancing in the waiting room, and reducing the number of providers and patients who were in the office at any one time.^1,2^ To accomplish the latter, they were asked to take a “virtual-first” approach and assess patients by phone, video, email or secure messaging before bringing them into the office.^3^ At the same time, many saw a dramatic drop in practice income due to total reduced visits in the first few months of the pandemic when patients were told to defer non-urgent care.^4,5^ Family physicians were also asked to support health system responses, for example, by staffing COVID19 assessment centres and helping in long-term care, overcrowded emergency departments and hospital wards, and later on by contributing to vaccination efforts.^5-7^

As a result of these dramatic changes, there have been concerns that the front door to our healthcare system was temporarily closed to a proportion of patients. Regulatory colleges indicate they have received complaints from patients of family physicians not seeing patients in-person months into the pandemic^8^ and some within the profession, particularly those staffing emergency rooms, have contended the same concern.^9^ Others have raised concerns that practices are closing altogether.^10-12^ However, there is little data to validate these anecdotal observations or understand the extent of these problems and the underlying reasons. In Canada, studies using administrative data have found that, one year into the pandemic, approximately 60% of primary care visits were virtual; ^13, 14^ it is unclear what portion of these are by phone versus video and there are no data on the portion that are by email or secure messaging. Patients seem to want virtual care to continue, ^15^ but it is unclear whether physicians agree and what supports physicians need to sustainably integrate virtual platforms into practice.

We conducted a survey of family physicians in Canada’s largest city to understand whether they remained open, especially to in-person visits, during the height of the second wave of COVID-19, possible reasons for closure, and associated physician and practice characteristics. We were also interested in family physician use of virtual care, desired virtual care supports, acceptance of new patients, and plans for future practice.

## Methods

### Setting and context

Toronto is Canada’s largest city with a population of 2.7 million.^16^ In 2015/16, Toronto had approximately 3500 primary care physicians, of which 2230 were thought to be providing comprehensive family medicine care.^17^ Approximately 80% of Toronto family physicians practice in a Patient Enrolment Model (PEM), a group of 3 or more physicians who have shared responsibility for after-hours access and receive some blended payments.^18^ Of those in an enrolment model, just over half are paid primarily by capitation with some incentives and fee-for-service income; the remaining are paid primarily fee-for-service with some incentives and only a small monthly capitation fee.^19^ Approximately 20% of capitation practices are part of a Family Health Team that receives funding for other health professionals like social workers and pharmacists and have added accountability for services provided. Finally, less than 2% of physicians practice in a Community Health Centre, a team-based, salaried model that traditionally serves more structurally marginalized communities. On March 14, 2020, new virtual billing codes were introduced in Ontario, compensating physicians for phone or video visits; no billing codes were introduced for email or secure messaging with patients.^20^ Phone and video codes were given the same fee-for-service dollar value as the analogous in-person visit billing codes and were considered “in-basket” within capitation models.

### Study Design and Population

Between March and June 2021, we conducted a cross-sectional survey of family physicians practicing in six geographic areas in Toronto recently aligned with Ontario Health Teams (Appendix 1). Because there is no single validated database of actively practicing family physicians in Ontario, we identified eligible family physicians using local knowledge and data from the regulatory college. First, family physician collaborators across these six geographic areas provided contact information to project staff for all family physicians who they believed were actively practicing office-based, comprehensive family medicine in their area. Second, this information was supplemented with publicly available information from the College of Physicians and Surgeons of Ontario (CPSO) gathered from a previous outreach initiative.^21^ We excluded physicians who did not practice office-based primary care in the 6 months prior to the pandemic (September 2019 to February 2020), had moved their practice outside of Toronto, were on parental leave, or were no longer practicing comprehensive family medicine (e.g. focused practice, practice closed, retired).

The project was initiated by family physician leaders in Toronto to directly inform regional policy and planning. It was sponsored by the regional health authority, Ontario Health Toronto Region, and supported by provincial partners including the Ontario College of Family Physicians and the Ontario Medical Association. The project was reviewed by institutional authorities at Unity Health Toronto and deemed to neither require Research Ethics Board approval nor written informed consent from participants.

### Survey

Three versions of a survey were developed, one for each of email, fax, and phone distribution (Appendix 2). The email survey was the most comprehensive and included questions on i) practice changes during the pandemic ii) practice operations in January 2021 including whether physicians were seeing patients in-person iii) virtual care, iv) future plans, and v) demographics as well as relevant probes (e.g. what factors influenced a decision not to be open to in-person visits). The fax included only a few questions from each of these areas with limited probes. The phone survey had the fewest questions and was designed to elicit responses from either the physician or their reception staff; it included key questions from sections i, ii, and v above with no probes. In cases where staff were unable to speak to a physician or receptionist, they noted relevant information from the practice voicemail greeting, when it was available.

The surveys were developed in collaboration with local and provincial family physician collaborators, a senior administrator at the regional health authority, and a survey methodologist. They were piloted by approximately ten practicing family physicians and revised accordingly. The email survey was electronic and hosted on Qualtrics, the fax survey was paper-based, and the phone survey was administered orally by trained project staff.

### Data Collection

The survey was first sent electronically to all family physicians for whom we had an email address. Physicians were sent a unique survey link and were sent up to three completion reminders over a 4 week period. Physicians who did not respond to the email survey, or for whom we had no email address, were sent the fax version of the survey; faxed surveys had a unique identifier corresponding to the relevant physician. We sent one fax reminder one week after the initial fax was sent. Physicians who did not respond to the email or fax surveys were then contacted by phone by trained project staff. After obtaining verbal consent, project staff asked questions of the reception staff, or were directed to speak directly with the physician in some cases. If there was no answer, staff left a voicemail and conducted one follow-up phone call one week after the initial phone outreach. In cases where staff were unable to speak to a physician or receptionist, they noted relevant information from the practice voicemail greeting, when it was available.

Email and fax survey introductions were signed by a local family physician collaborator. No financial incentive was provided for physicians to participate. Once data collection was complete, all personal identifiers were removed from the dataset and replaced with a study ID; physician names corresponding to the study ID were kept in a separate linking log.

### Analysis

We analyzed responses to surveys where at least one question was answered. Descriptive statistics were calculated; denominators were specified based on the number of physicians responding to a question. Chi-squared tests or Kruskal Wallis were used to test whether there was an association between physician and/or practice characteristics and whether the physician was i) seeing patients in-person, ii) seeing symptomatic patients, and iii) intended to close their practice in the next 5 years. A p-value ≤ 0.05 is considered statistically significant. All analyses were done in R version 4.0.0.

## Results

We identified 1339 family physicians who we thought met eligibility criteria based on publicly available information and information from our collaborators. Of these, 134 had an incorrect listing and an additional 19 responded to the survey indicating they did not provide office-based care in the 6 months before the pandemic and were thus excluded (Figure 1). We received and analyzed 1016 survey responses from the remaining 1186 eligible family physicians, 420 from email, 53 from fax, 390 from phone, and 153 from the voicemail greeting (overall response rate 86%). Those who responded to the survey had a more recent graduation year (Mean [SD]: 1998 [14.2] vs. 1994 [15.6], p<0.01) and a higher proportion were women (61.5% vs. 49.4%, p<0.01).

**Figure 1.**
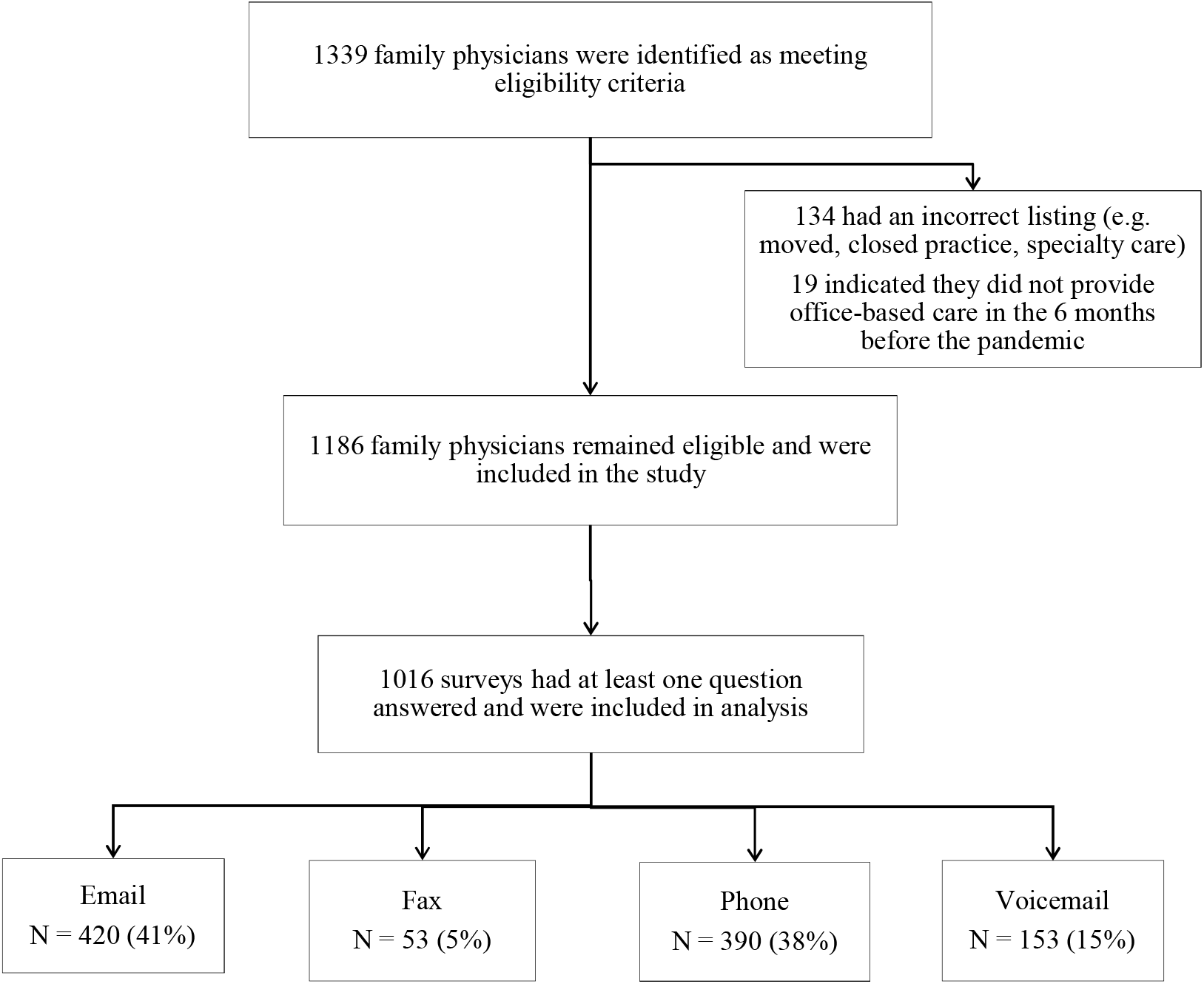
Flow diagram of family physicians eligible for and responding to the survey

**Figure 2.**
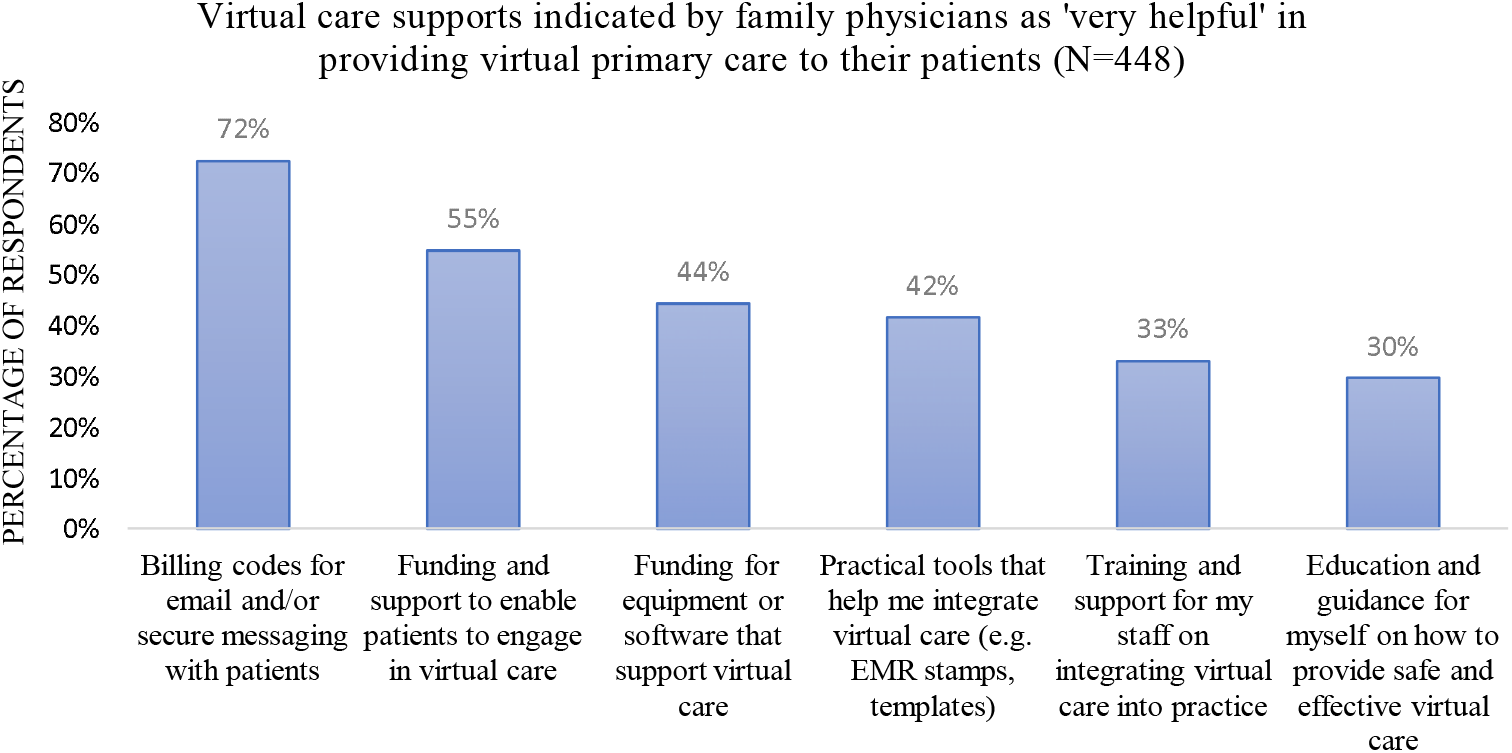
Virtual care supports indicated by family physicians as ‘very helpful’ in providing virtual primary care to their patients (N=448)

Respondents had a mean graduating year of 1998 and mean practice size of 1215 patients; 61.5% were women (Table 1). The majority worked in a group setting with only 12.8% reporting they were the only physician in their clinic. 46.9% worked in either a Family Health Team or Community Health Centre, 27.0% in a non-team capitation model, and 21.5% either in enhanced or straight fee-for-service. 2.7% of practices provided only walk-in services.

**Table 1.** Demographic characteristics of family physicians who responded to the survey (N=1016)

| Characteristic | N (%) |
| --- | --- |
| **Gender (n=1013)** |  |
| Woman | 623 (61.5%) |
| Man | 390 (38.5%) |
| **Practice remuneration model (n=755)** |  |
| PEM: Enhanced Fee-For-Service | 146 (19.3%) |
| PEM: Blended Capitation without team | 204 (27.0%) |
| PEM: Family Health Team | 308 (40.8%) |
| Community Health Centre | 46 (6.1%) |
| Traditional Fee-for-Service | 16 (2.1%) |
| Other | 35 (4.6%) |
| **Office practice setting (n=811)** |  |
| Group setting (2-5 physicians in my clinic) | 253 (31.2%) |
| Group setting (>5 physicians in my clinic) | 429 (52.9%) |
| Only physician in clinic | 104 (12.8%) |
| Works in multiple office settings | 25 (3.1%) |
| **Provides walk-in services only (n=778)** |  |
| Yes | 21 (2.7%) |
| No | 757 (97.3%) |
| **Approximate number of patients in practice (n=435)** |  |
| Mean (SD) | 1215 (901) |
| Median (IQR) | 1000 (775-1500) |
| **Year graduated from medical school (n=1013)** |  |
| Mean (SD) | 1998 (14.2) |
| Median (IQR) | 2001 (1986-2011) |

Note: Gender and year of graduation are from publicly available data from the College and Physicians and Surgeons of Ontario; other demographic variables are from respondent self-report. PEM=Patient Enrolment Model; Enhanced Fee-For-Service includes the Family Health Group and Comprehensive Care Model; Blended Capitation includes the Family Health Organization and Family Health Network.

### Whether the practice was open in January 2021

99.7% (1001/1004) of respondents indicated their practice was open to in-person or virtual visits in January 2021 with 94.8% (928/979) saying they saw patients in-person. Among those not seeing patients in-person, 100% (15/15) said they had arrangements for their patients to be assessed elsewhere with 60.0% reporting this was with another physician in their office or practice group. The most important factor for the decision not to see patients in-person was health concerns (93.7% or 15/16 reported as a fairly or very important factor) followed by supply of personal protective equipment (PPE) (26.7% or 4/15 reported as a fairly or very important factor). 60.0% (9/15) reported not seeing patients in-person for more than 6 months. Comparing physicians who did and did not see patients in-person, there were statistically significant differences in mean medical school graduation (mean [SD]: 1990 [18.1] among those seeing patients in-person vs 1999 [13.7] among those not seeing patients in-person, p<0.001) and office practice setting (among those seeing patients in-person, 12.4% were the only physician in their clinic vs 27.6% for those not seeing patients, p<0.05).

### Care of patients with COVID-19 symptoms

30.8% (264/856) of respondents said that in January 2021 they provided in-person care in their office to any patient reporting symptoms consistent with COVID-19. Among those who reported not seeing any symptomatic patient in-person, 59.3% (172/290) said they would refer symptomatic patients to the local testing centre and assess them in-person following a negative COVID-19 test while 33.8% (98/290) said they sent all symptomatic patients to the local emergency department or urgent care centre if in-person assessment was necessary. When comparing physicians who did and did not see symptomatic patients in-person, there was a statistically significant difference in mean estimated panel size (mean [SD]: 921 [671] vs 1328 [969], p<0.001), mean graduation year (mean [SD]: 2000 [12.7] vs 1996 [14.4], p<0.001), practice remuneration model (p<0.001), and office practice setting (p<0.001) (Table 2). Among those who saw symptomatic patients in-person, 69.5% were in a Family Health Team and 80.0% were in a group setting with >5 physicians compared to 27.3% and 40.8%, respectively, among those who did not see symptomatic patients.

**Table 2.** Characteristics of family physicians who did and did not report seeing patients with COVID-19 symptoms in their clinic in January 2021 (N= 856)

|  | Yes (N=264)<br>N (%) | No (N=592)<br>N (%) | P-value |
| --- | --- | --- | --- |
| **Gender (n=856)** |  |  |  |
| Male | 92 (34.8%) | 242 (40.9%) | 0.11 |
| Female | 172 (65.1%) | 350 (59.1%) |  |
| **Medical school graduation year (n=856)** |  |  |  |
| Mean (SD) | 2000 (12.7) | 1996 (14.4) | <0.001 |
| Median (IQR) | 2003 (1992-2011) | 1998 (1985-2010) |  |
| **Practice remuneration model (n=712)** |  |  |  |
| PEM: Enhanced Fee-For-Service | 21 (8.8%) | 118 (24.9%) | <0.001 |
| PEM: Blended Capitation without team | 22 (9.2%) | 169 (35.7%) |  |
| PEM: Family Health Team | 166 (69.5%) | 129 (27.3%) |  |
| Community Health Centre | 21 (8.8%) | 25 (5.3%) |  |
| Traditional Fee-for-Service | <6 (<2.5%) | 10 (2.1%) |  |
| Other | <6 (<2.5%) | 22 (4.6%) |  |
| **Office practice setting (n=765)** |  |  |  |
| A group setting (2-5 physicians in my clinic) | 20-35 (8.2%- 14.3%) | 214 (41.1%) | <0.001 |
| A group setting (>5 physicians in my clinic) | 196 (80.0%) | 212 (40.8%) |  |
| Only physician in clinic | 16 (6.5%) | 81 (15.6%) |  |
| Works in multiple office settings | <6 (<2.4%) | 13 (2.5%) |  |
| **Providing walk-in services only (n=732)** |  |  |  |
| Yes | <6 (<2.8%) | 16 (3.1%) | 0.52 |
| No | 195-210 (92.0%-99.1%) | 504 (96.9%) |  |
| **Estimated panel size (n=414)** |  |  |  |
| Mean (SD) | 921 (671) | 1328 (969) | <0.001 |
| Median (IQR) | 800 (500-1100) | 1100 (850-1550) |  |

Note: Gender and year of graduation are from publicly available data from the College and Physicians and Surgeons of Ontario; other demographic variables are from respondent self-report. PEM=Patient Enrolment Model; Enhanced Fee-For-Service includes the Family Health Group and Comprehensive Care Model; Blended Capitation includes the Family Health Organization and Family Health Network. Cell sizes <6 have been suppressed.

### Virtual care

Respondents estimated spending 27.2% of clinical care time in January 2021 doing in-person visits, 58.2% doing scheduled phone assessments, 5.8% doing scheduled video assessments, and 7.5% using secure messaging or email (Table 3). However, only 14.2% (64/450) and 8.2% (37/450) reported they were fairly or very likely to offer phone and video appointments, respectively, if virtual billing codes did not continue. In contrast, 93.6% (421/450) and 55.7% (250/449) said they were fairly or very likely to offer phone and video appointments, respectively, if the virtual billing codes continued. The most desired additional supports for virtual care were billing codes for email and/or secure messaging (72.3% [323/447] indicated this as very helpful) followed by funding and support to enable patients to engage in virtual care (54.7% [245/448] indicated this as very helpful).

**Table 3.**
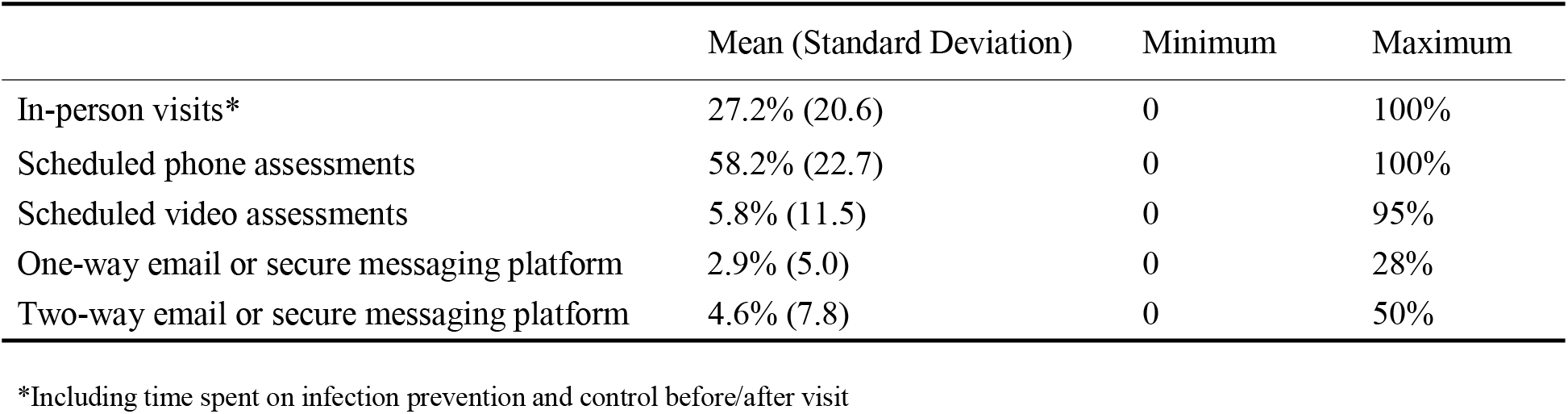
Estimated time spent by family physicians doing in-person and virtual care (N=450) Responses based on the question *“Think about all of the time you spent providing clinical care to patients in your office during January 2021. What portion of your time did you spend doing the following? (Please respond so that the total equals 100%)”*

### Acceptance of new patients, practice closure, and future practice intentions

Regarding new patients, 4.9% (22/448) said they were actively seeking to grow their practice and 11.6% (52/448) said they were accepting any new patients who contacted their office seeking care. 45.5% (204/448) said they only accepted family members of current patients while 38.0% (170/448) were not accepting any new patients.

Six physicians reported closing their practice permanently during the pandemic, some as previously planned and some earlier than planned. 3.4% (34/1016) reported hiring a locum to manage their patients while 2.4% (24/1016) reported temporarily closing their practice without locum coverage at some point from March 2020 to January 2021. The most commonly reported reason for temporarily hiring a locum was “needed a break” (43.5% [10/23] reported as very important).

At the time of the survey, 3.8% (17/447) of physicians were planning to close their current practice in the next year with an additional 13.4% (60/447) planning to close in the next 2-5 years. When comparing those who did and did not plan to close their practice in the next 5 years, there were statistically significant differences in medical school graduation year (mean [SD]: 1980 [8.8] vs 1998 [13.3], p<0.001), mean estimated panel size (mean [SD]: 1361 [809] vs 1195 [927], p<0.05), gender (p<0.01), office practice setting (p<0.01), and whether providing only walk-in services (p<0.05) (Table 4). Among those who planned to close their practice, 58.4% were men, 28.9% were the only physician in their clinic, and 7.9% were providing only walk-in services compared to 38.7%, 13.9%, and 2.2%, respectively, among those not planning to close their practice.

**Table 4.** Characteristics of family physicians who reported they were thinking of closing their practice in the next five years (N=439)

|  | Yes in the next 1 to 5 years<br>(N=77)<br>N (%) | No or not sure (N=362)<br>N (%) | P-value |
| --- | --- | --- | --- |
| **Gender (n=439)** |  |  |  |
| Male | 45 (58.4%) | 140 (38.7%) | <0.01 |
| Female | 32 (41.6%) | 222 (61.3%) |  |
| **Medical school graduation year (n=439)** |  |  |  |
| Mean (SD) | 1980 (8.8) | 1998 (13.3) | <0.001 |
| Median (IQR) | 1979 (1975-1985) | 1999 (1987-2009) |  |
| **Practice remuneration model (n=433)** |  |  |  |
| PEM: Enhanced Fee-For-Service | 25 (32.9%) | 76 (21.3%) | 0.34 |
| PEM: Blended Capitation without team | 28 (36.8%) | 137 (38.4%) |  |
| PEM: Family Health Team | 17 (22.4%) | 100 (28.0%) |  |
| Community Health Centre | <5 (<6.6%) | 12 (3.4%) |  |
| Traditional Fee-for-Service | <5 (<6.6%) | 13 (3.6%) |  |
| Other | <5 (<6.6%) | 19 (5.3%) |  |
| **Office practice setting (n=436)** |  |  |  |
| A group setting (2-5 physicians in my clinic) | 29 (38.2%) | 130 (36.1%) | <0.01 |
| A group setting (>5 physicians in my clinic) | 20-25 (26.3-32.9%) | 169 (46.9%) |  |
| Only physician in clinic | 22 (28.9%) | 50 (13.9%) |  |
| Works in multiple office settings | <6 (<7.9%) | 11 (3.1%) |  |
| **Providing walk-in services only (n=435)** |  |  |  |
| Yes | 6 (7.9%) | 8 (2.2%) | <0.05 |
| No | 70 (92.1%) | 351 (97.8%) |  |
| **Estimated panel size (n=423)** |  |  |  |
| Mean (SD) | 1361 (809) | 1195 (927) | <0.05 |
| Median (IQR) | 1200 (887-1600) | 1000 (750-1500) |  |

Note: Gender and year of graduation are from publicly available data from the College and Physicians and Surgeons of Ontario; other demographic variables are from respondent self-report. PEM=Patient Enrolment Model; Enhanced Fee-For-Service includes the Family Health Group and Comprehensive Care Model; Blended Capitation includes the Family Health Organization and Family Health Network. Cell sizes <6 have been suppressed.

## Discussion

Our survey of Toronto-area family physicians found that 99.7% of practices remained open and 94.8% were seeing at least some patients in-person in January 2021, during the height of the second wave of COVID-19 in Ontario, ^22^ prior to the widespread availability of vaccinations. Personal health concerns was the most common reason physicians identified for not seeing any patients in-person; all physicians who reported not seeing patients in-person in January 2021 had arrangements for their patients to be seen by a colleague if needed. Less than a third of physicians reported seeing any patient with COVID-19 symptoms in their office. However, among the two-thirds who reported not seeing symptomatic patients in-person, 59.3% said they would do so after the patient received a negative COVID-19 test. A higher portion of those seeing symptomatic patients reported being part of a team-based practice and working in a clinic with more than 5 other physicians. Almost one in five physicians reported thinking about closing their practice in the next 5 years; a higher proportion were men with an older graduation year who reported being the only physician in their clinic.

Our findings run counter to a popular narrative that family physician offices were closed during COVID-19.^23^ However, our results raise concerns about a shrinking workforce with a substantial number of physicians considering closing practice in the near future—a worrisome finding in the context of one in ten Canadians already reporting not having a regular family physician.^24^ Our results are in keeping with other research that has suggested that the pandemic has caused some family physicians to stop working and potentially accelerating retirement plans.^10,25,26^ Our study did not explore reasons for wanting to leave practice in the next 5 years but possible hypotheses include health concerns, financial issues, and burnout.^12,27-29^Physicians in our study reported that in January 2021 almost two-thirds of care was delivered virtually, the vast majority by phone. Most physicians would like to continue to provide scheduled, appointment-based phone care post-pandemic, but only if virtual billing codes continue. The most desired support for virtual care was for billing codes for email or secure messaging suggesting physicians see value in integrating these into clinical care despite our finding that only 7.5% of clinical time was used on these modalities. Previous studies have also found that secure messaging was popular for both patients and physicians^30^ and physician remuneration was a potential facilitator for increased uptake.^31^ Physicians in our study also wanted support for patients to engage in virtual care which aligns with research that has found virtual care leaves some groups of patients behind.^32-34^

COVID-19 has also highlighted the variation in primary care infrastructure and accountability. In our study, more physicians who worked alone in a clinic reported not seeing patients in-person, not seeing symptomatic patients, or considering closing their practice in the next 5 years compared to physicians who worked in groups with more than 5 physicians. These findings highlight the particular challenges of traditional, fee-for-service solo practice in the pandemic and are in keeping with calls for payment and organizational reform in primary care.^35-37^ Team-based practices in Ontario have dedicated administrative support as well as formal accountabilities including reporting for timely access,^38^ which may explain why more physicians in these models reported seeing symptomatic patients in clinic. We also hypothesize that team-based and group practices have larger waiting rooms, allowing for more patients to be safely seen in-person.

Our study has strengths and limitations. We conducted a systematic survey of all family physicians practicing in 6 geographic areas in Toronto and achieved an 86% response rate on our core questions of whether or not a practice was open and seeing patients in-person. However, our sample is open to non-response bias; it is possible that those who did not respond were more likely to be closed or not seeing patients in-person. As well, family physicians self-reported on whether they were open or closed and may have been reluctant to disclose their practice closure. Physician self-report may also have been different from patient perceptions of whether a practice was open; the latter may be influenced by longer wait times during the pandemic, but our study was not designed to assess this. Despite our extensive outreach and high response rate, we had fewer respondents working in enhanced or straight fee-for-service models relative to population distribution. Neighbourhoods in Toronto were differently affected by COVID-19^39^ and our survey did not explore the impact of neighbourhood context on family physician decisions. Results may also not be generalizable to other contexts. Finally, we asked respondents to reflect on practice patterns 2-5 months prior to the time of the survey which may have influenced accuracy of recall. It is also worth noting that there is no Canadian data we are aware of on how specialist physician practices responded to the pandemic.

### Conclusion

Our survey found that the vast majority of family physician practices in Canada’s largest city were open and seeing patients in-person during COVID-19—even before widespread vaccination of health care workers and the general population. Our findings contrast with media stories of patients reporting their family physicians were not seeing patients—an important perspective that warrants further study. Our findings also highlight the challenge of operating a solo family practice during the pandemic and support calls to expand group practice opportunities and access to team-based models that include administrative support and accountability—policy directions that may also influence more medical graduates to choose family medicine as a career.^36,40^ Recruiting and retaining the primary care workforce is particularly important given that one in five physicians in our study said they were thinking about closing their practice in the next five years. Understanding and addressing root causes of burnout will be important to prevent physicians from exiting practice. Finally, to integrate virtual care into routine practice post-pandemic, we need to consider appropriate financial remuneration for physicians—including for email or secure-messaging—as well as supports for patients who struggle with virtual connectivity.

## Supporting information

Appendix 1- KDO Study Regions

Appendix 2- Keeping Doors Open (KDO) Survey Questions

## Data Availability

Following peer-reviewed publication, de-identified data can be made available on request for proposed projects advancing the public good.

## Acknowledgements

Thank you to primary care collaborators in Toronto who co-led this effort including David Kaplan, Tia Pham, Art Kushner, Karen Fleming, David Schieck, Leanne Clark, Elizabeth Muggah, Allan Grill, staff who supported the outreach, and other members of the project working group including Dorothy Wedel and Valeria Rodriguez. Thank you to members of the MAP Survey Research Unit at St. Michael’s Hospital who also supported outreach including Alexandra Carasco, Natalie Johnson, Annika Khan, and Olivia Spandier. Thanks to Raphael Goldman-Pham, Hilarie Stein, and Jennifer Truong who supported an earlier outreach effort that informed this work as well as Adam Cadotte for help identifying physicians practicing in Toronto, Kirsten Eldridge for administrative support, and Amy Craig-Neil for initial coordination support.

## Notes

**Funding statement** This project received funding from INSPIRE Primary Health Care Research Program which is funded through the Health Systems Research Program of the Ontario Ministry of Health. The opinions, results and conclusions reported in this paper are those of the authors and are independent from the funding sources. No endorsement by the Ontario MOH is intended or should be inferred. Dr. Kiran is the Fidani Chair of Improvement and Innovation in Family Medicine at the University of Toronto and is also supported as a Clinician Scientist by the Department of Family and Community Medicine (DFCM) at the University of Toronto and at St. Michael’s Hospital.

**Conflicts of interest** The authors declare no conflicts of interest.

### Competing Interest Statement

The authors have declared no competing interest.

### Funding Statement

This project received funding from INSPIRE Primary Health Care Research Program which is funded through the Health Systems Research Program of the Ontario Ministry of Health. The opinions, results and conclusions reported in this paper are those of the authors and are independent from the funding sources. No endorsement by the Ontario MOH is intended or should be inferred. Dr. Kiran is the Fidani Chair of Improvement and Innovation in Family Medicine at the University of Toronto and is also supported as a Clinician Scientist by the Department of Family and Community Medicine (DFCM) at the University of Toronto and at St. Michaels Hospital.

### Author Declarations

The project was reviewed by institutional authorities at Unity Health Toronto and deemed to neither require Research Ethics Board approval nor written informed consent from participants.

