## Appendix 1- KDO Study Regions for "Keeping doors open: A cross-sectional survey of family physician practice patterns during COVID-19, needs, and intentions"

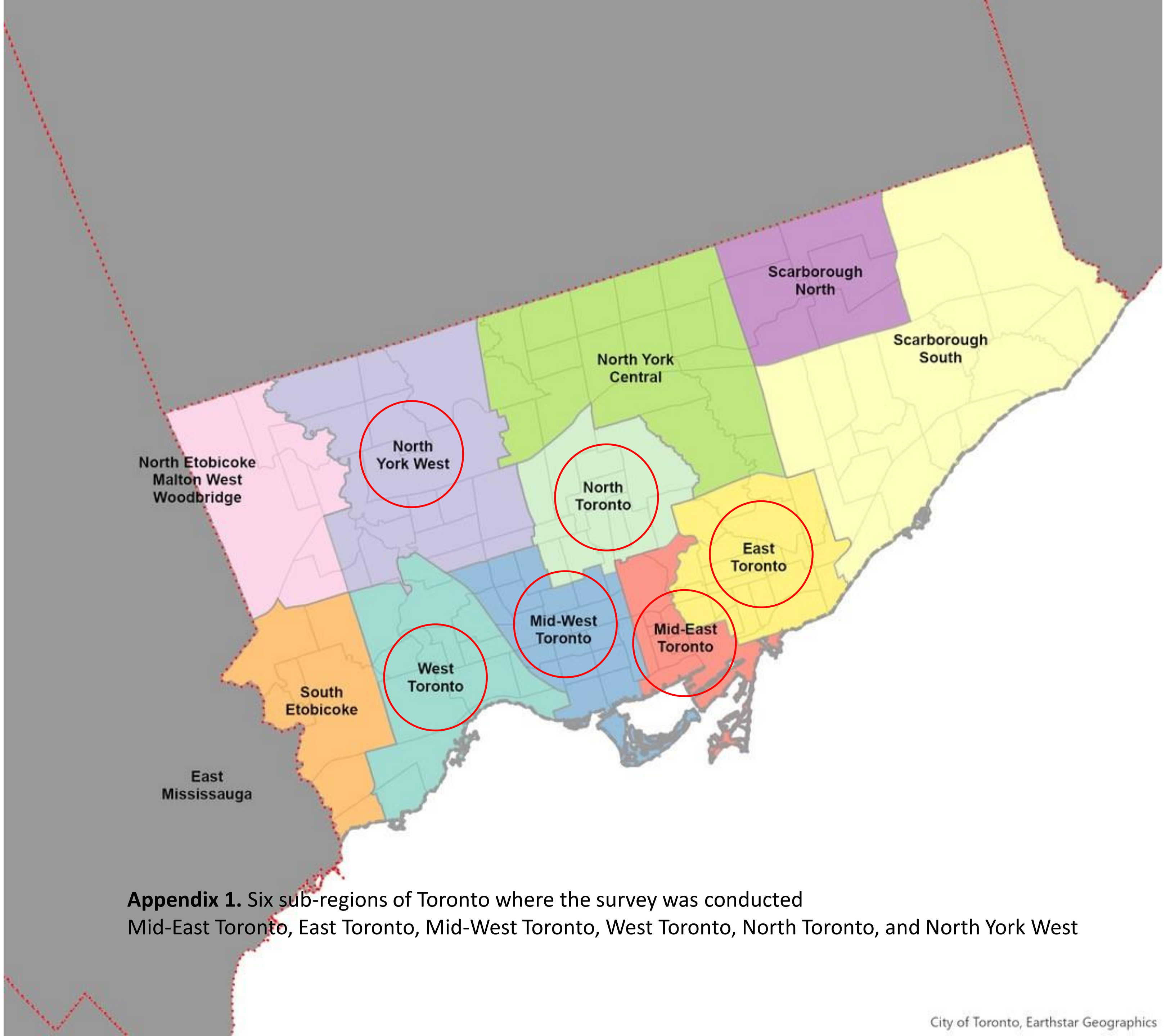


**Appendix 1.** Six sub-regions of Toronto where the survey was conducted
Mid-East Toronto, East Toronto, Mid-West Toronto, West Toronto, North Toronto, and North York West
