## Appendix 2- Keeping Doors Open (KDO) Survey Questions for "Keeping doors open: A cross-sectional survey of family physician practice patterns during COVID-19, needs, and intentions"

Note: All of the following questions were asked in the email survey in the order in which they appear in the table; columns indicate whether the question was included in other modes and the corresponding question number.

| **Survey Question** | **Fax** | **Telephone** | **VM Greeting** |
| --- | --- | --- | --- |
| **Section 1: Your clinical work before the pandemic** | | | |
| Did you provide any office-based primary care in the six months before the pandemic (Sept. 2019 to Feb. 2020)? | Q1 | Q1 | --- |
| What clinical activities did you engage in during the six months pre-pandemic?  Q4_1 Emergency Department  Q4_2 Focused practice (e.g. sports medicine, psychotherapy, pain medicine)  Q4_3 Long-term care  Q4_4 Outpatient rehabilitation  Q4_5 Hospitalist work  Q4_6 Clinical care in shelters  Q4_7 I do not do clinical work  Q4_8 Other (please specify) | --- | --- | --- |
| **Section 2: Change in practice operation after the pandemic began** | | | |
| Since the start of pandemic March 2020, have you done any of the following?  Q7_1 Temporarily closed practice without locum coverage  Q7_2 Hired a locum to manage your patients  Q7_3 Decided to close your practice permanently, earlier than planned  Q7_4 Decided to close your practice permanently, as previously planned | Q2 | Q2 | --- |
| How important were the following factors in your decision to close your practice temporarily?  Q8_1 Worried about personally contracting COVID-19  Q8_2 Not enough volume/revenue to continue operating practice  Q8_3 Unable to retain support staff  Q8_4 Other clinical work opportunities or re-deployment  Q8_5 Other non-clinical work or projects  Q8_6 Needed a break  Q8_7 Maternity/parental leave  Q8_8 Personal, health, or family reasons requiring time away from work  Q8_9 Other (please specify) | --- | --- | --- |

| **Survey Question** | **Fax** | **Telephone** | **VM Greeting** |
| --- | --- | --- | --- |
| How important were the following in your decision to temporarily hire a locum?  Q9_1 Worried about personally contracting COVID-19  Q9_2 Not enough volume/revenue to continue operating practice  Q9_3 Unable to retain support staff  Q9_4 Other clinical work opportunities or re-deployment  Q9_5 Other non-clinical work or projects  Q9_6 Needed a break  Q9_7 Maternity/parental leave  Q9_8 Personal, health, or family reasons requiring time away from work  Q9_9 Other (please specify) | --- | --- | --- |
| How important were the following factors in your decision to close your practice permanently, earlier than planned?  Q10_1 Worried about personally contracting COVID-19  Q10_2 Not enough volume/revenue to continue operating practice  Q10_3 Unable to retain support staff  Q10_4 Other clinical work opportunities or re-deployment  Q10_5 Other non-clinical work or projects  Q10_6 Needed a break  Q10_7 Maternity/parental leave  Q10_8 Personal, health, or family reasons requiring time away from work  Q10_9 Other (please specify) | --- | --- | --- |
| **Section 3: Current practice operations** | | | |
| In January 2021, was your practice open to in-person and/or virtual visits? | Q3 | Q3 | Q3 |
| How important were each of the following factors in your decision to close your practice?  Q13_1 Health concerns  Q13_2 PPE supply  Q13_3 PPE cost  Q13_4 Cost of environmental cleaning  Q13_5 Loss of income  Q13_6 Needed a break  Q13_7 Redeployment  Q13_8 Other (please specify) | --- | --- | --- |

| **Survey Question** | **Fax** | **Telephone** | **VM Greeting** |
| --- | --- | --- | --- |
| How long has your practice been closed? | --- | --- | --- |
| While your practice was closed, did you have arrangements for your primary care patients to get primary care elsewhere? | --- | --- | --- |
| In January 2021, did you see any patients in-person? | Q4 | Q4 | Q4 |
| How important were each of the following factors in your decision NOT to see patients in person?  Q17_1 Health concerns  Q17_2 PPE supply  Q17_3 PPE cost  Q17_4 Cost of environmental cleaning  Q17_5 Loss of income  Q17_6 Needed a break  Q17_7 Redeployment  Q17_8 Other (please specify) | Q5 | --- | --- |
| How long has your practice NOT seen patients in person? | --- | --- | --- |
| While your practice was NOT seeing patients in person, did you have arrangements for your primary care patients to get primary care elsewhere? | --- | --- | --- |
| Thinking about January 2021, did you provide in-person care in your office to patients reporting symptoms consistent with COVID-19 (for example, patients with shortness of breath, cough, or fever)? | Q6 | Q5 | Q5 |
| During the month of January 2021, how did you typically care for patients who reported symptoms consistent with COVID-19 who did not have an emergency but needed an in-person assessment? | --- | --- | --- |
| Think about all of the time you spent providing clinical care to patients in your office during January 2021. What portion of your time did you spend doing the following?  Q22_1 In-person visits (including time spent on IPAC before/after)  Q22_2 Scheduled phone assessments  Q22_3 Scheduled video assessments  Q22_4 One-way e-mail or secure messaging platform (provider can initiate message to patient but patient cannot initiate message)  Q22_5 Two-way e-mail or secure messaging platform (either patient or provider can initiate message exchange)  Q22_6 Other (please specify) | Q7 | --- | --- |
| Thinking about January 2021, were you accepting new patients for primary care? | Q8 | --- | --- |

| **Survey Question** | **Fax** | **Telephone** | **VM Greeting** |
| --- | --- | --- | --- |
| **Section 4: Virtual care** | | | |
| Consider the following scenarios with respect to virtual billing codes (K080, K081, K082):  Q25_1 If the virtual billing codes remain in place after the pandemic ends, will you offer **phone** appointments to your patients?  Q25_2 If the virtual billing codes remain in place after the pandemic ends, will you offer **video** appointments to your patients?  Q25_3 If the virtual billing codes do NOT remain in place after the pandemic ends, will you offer **phone** appointments to your patients?  Q25_4 If the virtual billing codes do NOT remain in place after the pandemic ends, will you offer **video** appointments to your patients? | Q9 | --- | --- |
| How helpful would each of the following supports be for you in providing virtual primary care to your patients?  Q26_1 Funding for equipment or software  Q26_2 Billing codes for email and/or secure messaging with patients  Q26_3 Education and guidance for myself on how to provide safe and effective virtual care  Q26_4 Training and support for my staff on integrating virtual care into practice  Q26_5 Practical tools that help me integrate virtual care (e.g. EMR stamps, templates)  Q26_6 Funding and support to enable patients to engage in virtual care | Q10 | --- | --- |
| **Section 5: Future Plans** | | | |
| Do you plan to close your current practice in the next 5 years? | Q11 | --- | --- |
| **Demographics** | | | |
| What is your practice remuneration model? | Q14 | Q6 | --- |
| What is your office practice setting? | Q15 | Q7 | --- |
| Do you provide walk-in services only? | Q16 | Q8 | --- |
| Approximately how many patients are in your practice? | Q17 | --- | --- |
| What is your gender? | Q12 | --- | --- |
| What year did you graduate from medical school? | Q13 | --- | --- |

Note: the question numbers do not always appear in order. This is due to the order in which questions were programmed in Qualtrics (survey software).

**Table A2** List of questions used in the KDO survey with sub-questions removed for easier viewing (abbreviated list)

|  | **Q# in Other Modes (if applicable)** | | |
| --- | --- | --- | --- |
| **Text** | **Fax** | **Telephone** | **VM Greeting** |
| Did you provide any office-based primary care in the six months before the pandemic (Sept. 2019 to Feb. 2020)? | Q1 | Q1 | --- |
| What clinical activities did you engage in during the six months pre-pandemic? | --- | --- | --- |
| Since the start of pandemic March 2020, have you done any of the following? | Q2 | Q2 | --- |
| How important were the following factors in your decision to close your practice temporarily? | --- | --- | --- |
| How important were the following in your decision to temporarily hire a locum? | --- | --- | --- |
| How important were the following factors in your decision to close your practice permanently, earlier than planned? | --- | --- | --- |
| In January 2021, was your practice open to in-person and/or virtual visits? | Q3 | Q3 | Q3 |
| How important were each of the following factors in your decision to close your practice? | --- | --- | --- |
| How long has your practice been closed? | --- | --- |  |
| While your practice was closed, did you have arrangements for your primary care patients to get primary care elsewhere? | --- | --- |  |
| In January 2021, did you see any patients in-person? | Q4 | Q4 | Q4 |
| How important were each of the following factors in your decision NOT to see patients in person? | Q5 | --- | --- |
| How long has your practice NOT seen patients in person? | --- | --- | --- |
| While your practice was NOT seeing patients in person, did you have arrangements for your primary care patients to get primary care elsewhere? | --- | --- | --- |
| Thinking about January 2021, did you provide in-person care in your office to patients reporting symptoms consistent with COVID-19 (for example, patients with shortness of breath, cough, or fever)? | Q6 | Q5 | Q5 |
| During the month of January 2021, how did you typically care for patients who reported symptoms consistent with COVID-19 who did not have an emergency but needed an in-person assessment? | --- | --- | --- |
| Think about all of the time you spent providing clinical care to patients in your office during January 2021. What portion of your time did you spend doing the following? | Q7 | --- | --- |
| Thinking about January 2021, were you accepting new patients for primary care? | Q8 | --- | --- |
| Consider the following scenarios with respect to virtual billing codes (K080, K081, K082) | Q9 | --- | --- |
| How helpful would each of the following supports be for you in providing virtual primary care to your patients? | Q10 | --- | --- |
| Do you plan to close your current practice in the next 5 years? | Q11 | --- | --- |
| What is your practice remuneration model? | Q14 | Q6 | Q6 |
| What is your office practice setting? | Q15 | Q7 | --- |
| Do you provide walk-in services only? | Q16 | Q8 | --- |
| Approximately how many patients are in your practice? | Q17 | --- | --- |
| What is your gender? | Q12 | --- | --- |
| What year did you graduate from medical school? | Q13 | --- | --- |
